# Can Deep Learning Models Differentiate Atrial Fibrillation from Atrial Flutter?

**DOI:** 10.1101/2023.08.08.23293815

**Authors:** Estela Ribeiro, Quenaz Bezerra Soares, Felipe Meneguitti Dias, Jose Eduardo Krieger, Marco Antonio Gutierrez

## Abstract

Atrial Fibrillation (AFib) and Atrial Flutter (AFlut) are prevalent irregular heart rhythms that poses significant risks, particularly for the elderly. While automated detection systems show promise, misdiagnoses are common due to symptom similarities. This study investigates the differentiation of AFib from AFlut using standard 12-lead ECGs from the PhysioNet CinC Challenge 2021 (CinC2021) databases, along with data from a private database. We employed both one dimensional-based (1D) and image-based (2D) Deep Learning models, comparing different 1D and 2D Convolutional Neural Network (CNN) architectures for classification. For 1D models, LiteVGG-11 demonstrated the highest performed, achieving an accuracy (Acc) of 77.91 (±1.73%), area under the receiver operating characteristic curve (AUROC) of 87.17 (±1.29%), F1 score of 76.59 (±1.90%), specificity (Spe) of 71.69 (±4.73%), and sensitivity (Se) of 86.53 (±5.33%). On the other hand, for 2D models the EfficientNet-B2 outperformed other architectures, with an Acc of 75.20 (±3.38%), AUROC of 85.50 (±1.14%), F1 of 71.59 (±3.66%), Spe of 74.76 (±13.85%) and Se of 75.74 (±13.85%). Our findings indicate that distinguishing between AFib and AFlut is non-trivial, with 1D signals exhibiting superior performance compared to their 2D counterparts. Furthermore, it’s noteworthy that the performance of our models on the CinC2021 databases was considerably lower than on our private dataset.

## I. INTRODUCTION

Atrial Fibrillation (AFib) and Atrial Flutter (AFlut) are distinct irregular heart rhythms originating from abnormal activity in the heart’s upper chambers, the Atria [1], [2]. These conditions pose significant risks, especially for the elderly [3]. AFib involves chaotic electrical activity, causing rapid, irregular atrial contractions at 350-500 beats per minute, compromising heart function and raising stroke risk [4], [5]. AFlut, often misdiagnosed as AFib, features a single electrical circuit driving atrial contractions at 250-350 beats per minute, disrupting heart function [4]. Early diagnosis and treatment are crucial for managing AFib and AFlut and reducing severe complications such as stroke [3].

Subtle or absent symptoms often accompany irregular heart rhythms, including chest pain, dizziness, shortness of breath, fainting, and palpitations [3], [5], associated with rapid ventricular rate and inadequate diastolic ventricular filling [2]. Automated detection systems can significantly aid in promptly and accurately identifying these conditions, improving healthcare efficiency and reducing patient wait times. This is especially beneficial for underprivileged hospitals with limited access to experienced cardiologists, alleviating strain on their healthcare infrastructure.

The electrocardiogram (ECG), an essential tool for diagnosing cardiac issues, is utilized extensively worldwide, with millions of exams conducted annually. ECG involves the measurement of the heart’s electrical activity using electrodes affixed to patient’s skin and is considered the gold standard for noninvasive diagnosis of various heart disorders [4]. Clinical assessment of AFib and AFlut predominantly relies on non-invasive 12-lead ECGs where distinct patterns of electrical activity on the ECG signal enable differentiation between these two conditions [1], [5].

On the ECG, AFib is characterized by the absence of P waves, irregular RR intervals, and fibrillatory waves, while AFlut typically displays sawtooth flutter waves [4]. However, despite these distinct patterns, AFlut is often misdiagnosed as AFib due to similar symptoms and AFib’s higher prevalence [2], [3], [5]. Some studies suggest that AFlut may be misinterpreted as AFib, especially when ventricular activity is highly irregular, causing AFlut to mimic AFib on surface ECGs [6]. This misinterpretation can lead to inappropriate treatment, as each condition requires a specific therapeutic approach.

Over the past decades, the research community has increasingly focused on automating AFib detection, with Deep Learning (DL) emerging as an effective technique for ECG analysis [7], [8]. Studies consistently show high accuracy in detecting AFib compared to non-AFib classes [9]–[13], with some proposing merging AFib and AFlut into a single class for classification [12]. However, distinguishing between AFib and AFlut has received limited attention, and existing studies have produced unsatisfactory results [9].

Most studies differentiating between AFib and AFlut typically use datasets such as the MIT-BIH Atrial Fibrillation [14], [15] and MIT-BIH Arrhythmia [15], [16], featuring extended records of two-lead one-dimensional ECGs. These studies adopt a classification approach for ECG signals, categorizing them into AFib, AFlut, and Normal Sinus Rhythm, often with limited subject pools. Consequently, they frequently had to partition this data into smaller segments for analysis. While employing the same subject for both training and testing sets may yield more precise results due to intra-subject heartbeat interdependence, caution is necessary, as relying solely on intra-subject paradigms could lead to overly optimistic and biased classifications [17]–[19].

Furthermore, most studies classifying ECGs rely on one-dimensional signals [13]. However, in clinical practice, physicians diagnose by visually examining and interpreting 12-lead ECGs exams. Thus, we hypothesize that bi-dimensional (image-based 12 lead ECG exams) DL models designed for AFib and AFlut discrimination may outperform one-dimensional models. Additionally, considering the common occurrence of misdiagnoses between these conditions, we also anticipate sub-optimal performance from DL models.

In this study, our aim is to investigate the effectiveness of employing 12-lead ECGs to differentiate between AFib and AFlut, utilizing either one-dimensional signals or traditional 12-lead ECG images, with a binary classification approach. We utilized data from the six largest PhysioNet Cinc Challenge 2021 (CinC2021) databases, along with a private database sourced from ambulatory patients at a tertiary referral hospital. We explored two types of input data: images (2D) and one-dimensional (1D) signals, with the objective of determining which yields better performance. This approach distinguishes our study from other state-of-the art DL-based ECG classification research. To conduct our experiments, we evaluated the performance of various different Convolutional Neural Network (CNN) architectures for image-based and one-dimensional-based input data. To the best of our knowledge, this study represent the first report on the assessment of AFib and AFlut discrimination using end-to-end CNNs. Our study offers the following contributions:

1. Thorough evaluation of CNN models for distinguishing between AFib and AFlut based on ECG data;
2. Comparative analysis of 1D and image-based CNN models, shedding light on their relative efficacy in arrhythmia classification tasks;
3. Analysis of different datasets highlighting the importance of dataset composition and balance in model performance.

## II. METHODS

In this section, we describe the dataset, the preprocessing steps and the deep neural networks architecture used for binary classification of ECG signals. The general structure of the proposed method is shown in Figure 1.

**FIGURE 1.**
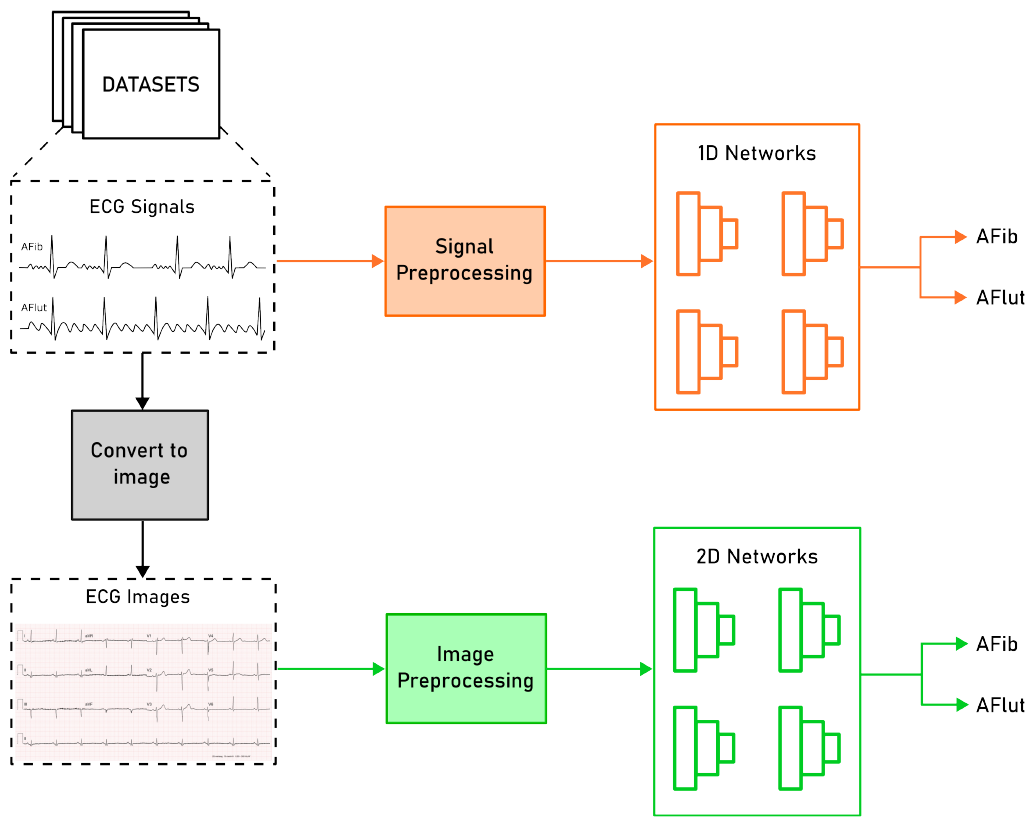
General structure of the proposed methodology for AFib and AFlut classification based on ECG data with different 1D and 2D CNN architectures.

All of our experiments were performed using a Foxconn High-Performance Computer (HPC) M100-NHI with an 8 GPU cluster of 32 GB NVIDIA Tesla V100 cards. The methodology was implemented using the Python framework (version 3.6.8) and Keras/TensorFlow (version 2.3.0).

### A. DATASETS

We utilized the PhysioNet CinC Challenge 2021 (CinC2021) databases [20], [21], which offer a repository of standard 12-lead ECGs covering 30 cardiac abnormality diagnoses. It comprises the following datasets: public (CPSC) and unused (CPSC extra) China Physiological Signal Challenge, St. Petersburg Institute of Cardiological Technics (INCART), Physikalisch-Technische Bundesanstalt (PTB), PTB-XL, Georgia 12-lead ECG Challenge, Chapman Shaoxing and Ningbo.

Typically, 12-lead ECGs present 10 s of recorded signals. To avoid losing arrhythmic morphologies present in long-term ECG labels, we selected datasets with recordings of approximately 10 s. Therefore, we excluded PTB and INCART datasets due to longer recordings exceeding 10 s.

We also incorporated a private dataset of 12-lead ECGs, denoted as InCor-DB, comprising data collected between 2017 and 2020 [13]. This dataset was sourced from the Picture Archiving and Communication System (PACS) of a specialized tertiary referral hospital in Brazil with focus on cardiology, namely Heart Institute Hospital. Data were acquired using MORTARA TM ELI 250c machines, encompassing 52 distinct clinical diagnoses related to cardiac abnormalities. It is important to note that this private dataset fully adheres to all pertinent ethical regulations and was approved from the Institutional Review Board (IRB).

This study aimed to analyze patient data diagnosed with AFib and AFlut arrhythmia, excluding records with different diagnostic annotations. We utilized class weight estimation techniques to handle dataset imbalance. Table 1 outlines the number of ECGs in the six largest CinC2021 databases and the InCor-DB dataset.

**TABLE 1.** Number of selected 12-lead ECG exams from the six CinC2021 datasets and the InCor-DB private database.

| Datasets | ECGs (Total) | Atrial Fibrillation | Atrial Flutter |
| --- | --- | --- | --- |
| Chapman-Shaoxing 12-lead ECG | 2,225 | 1,780 | 445 |
| CPSC 2018 Training Set (CPSC 2018) | 1,221 | 1,221 | 0 |
| China 12-Lead ECG (CPSC2018-Extra) | 207 | 153 | 54 |
| Georgia 12-Lead ECG Challenge | 756 | 570 | 186 |
| Ningbo First Hospital 12-lead ECG | 7,615 | 0 | 7,615 |
| PTB-XL Electrocardiography | 1,587 | 1,514 | 73 |
| Private InCor-DB 12-Lead ECG | 9,528 | 8,219 | 1,309 |
| *Total* | *23,139* | *13,457* | *9,682* |

### B. DATA PREPROCESSING

The standard 12-lead ECG raw signals were resampled to 500 Hz and standardized to a length of 10 seconds. This entailed either truncating longer signals to the initial 10 seconds or zero-padding shorter signals to achieve the desired duration. Our preprocessing consists of two phases: one for 1D signals and the other for 2D signals, which are essentially images. Figure 2 displays our preprocessing approach.

**FIGURE 2.**
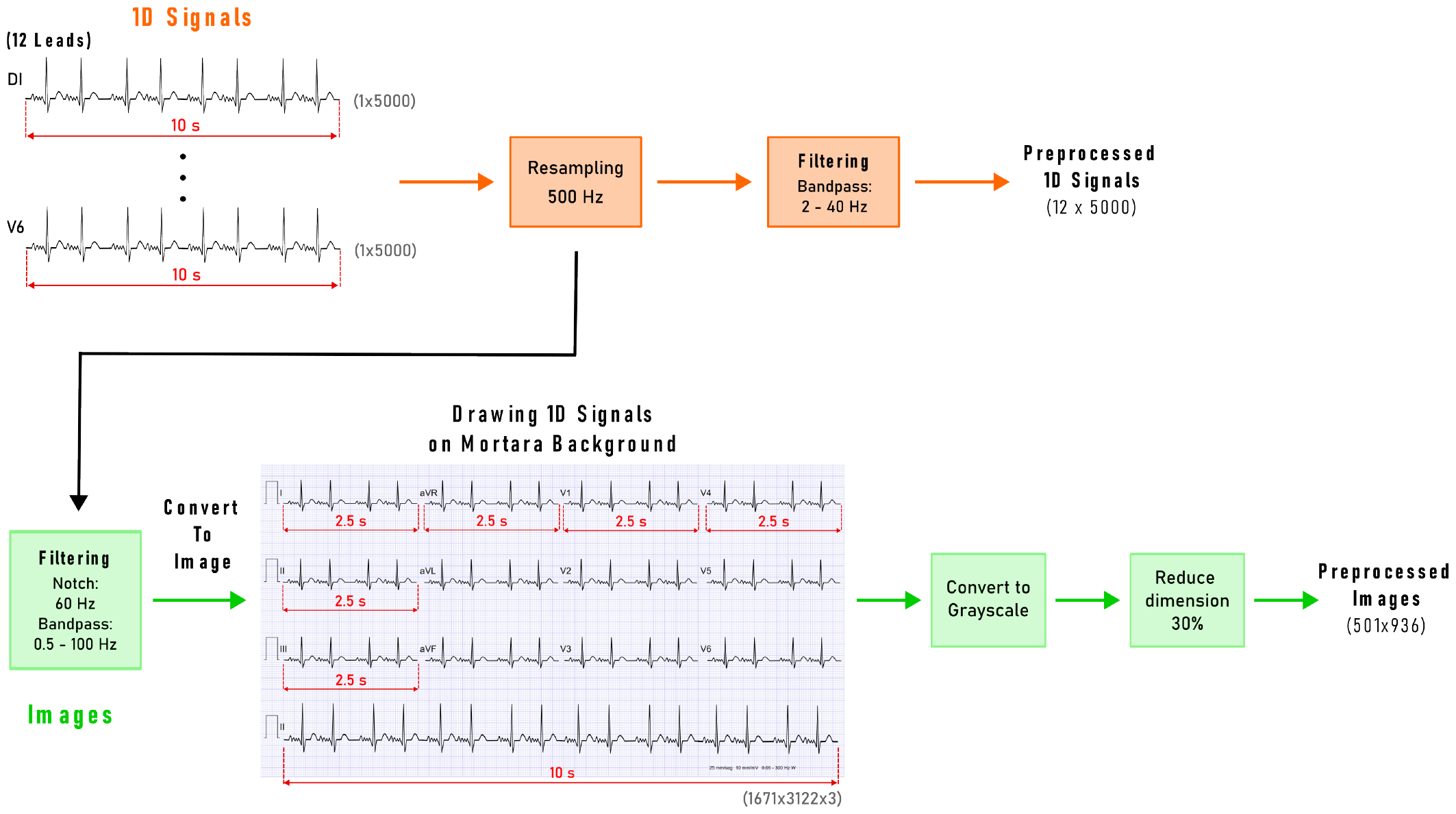
Preprocessing Steps for 1D and 2D ECG signals.

For 1D signals, we applied a Butterworth bandpass filter with a frequency range of 2-40 Hz, maintaining the original sampling rate of 500 Hz.

To create the dataset for 2D signals, we converted the 1D raw signals from the original datasets into images using the MORTARA ECG image template, with the signals drawn onto this background. The original image dimensions were 1671×3122×3. We opted for the MORTARA template to mimic how physicians would typically encounter ECG exams. Prior to conversion, the signals underwent filtering with a 60 Hz notch filter and a 0.5–100 Hz bandpass Butterworth filter. We then converted the images to grayscale and resized them to 30% of their original dimensions (resulting in a 501×936 grayscale image) to reduce computational complexity.

### C. DEEP LEARNING MODELS

#### 1) One-dimensional Classification

We employed seven 1D CNN architectures to assess the performance of AFib and AFlut classification in 1D data: (i) LiteVGG-11 [22]; (ii) LiteResNet-18 [22]; (iii) MobileNet [23]; (iv) ResNet-50 [24]; (v) VGG-16 [25]; (vi) DenseNet-121 [26]; and (vii) EfficientNet-B2 [27].

For the traditional CNNs, we adapted the 2D convolutions to 1D convolutions. In the case of Lite models (LiteVGG-11 and LiteResNet-18), we implemented a lightweight CNN proposed by Quenaz et al. (2022) [22]. These Lite models deliver comparable performance to their original counterparts, while demanding fewer computing resources. Their approach incorporates depth-wise separable 1D convolution layers (DWConv), a reduced number of filters, a global average pooling for flattening, and fewer units in the dense layers. We retained the fully connected layers of the original models, only modifying the replacement of the last layer with a single output using a sigmoid activation. Each model underwent training for over up to 120 epochs with a batch size of 64. To mitigate overfitting, we incorporated an early stopping callback with patience of seven epochs. This means that if the model does not improve in the validation dataset for seven consecutive epochs, the training process is stopped.

#### 2) Image Classification

To assess the performance of image-based classification of AFib and AFlut, we used five traditional and widely used 2D CNNs: (i) MobileNet [23]; (ii) ResNet-50 [24]; (iii) VGG16 [25]; (iv) Densenet-121 [26]; and (v) EfficientNetB2 [27].

The fully connected layers comprised a customized 3-layer perceptron with dropout regularization set at 30%, ReLU activation function in intermediate layers, and a sigmoid function in the final layer. Each model underwent training for 30 epochs, utilizing a batch size of 8. Similar to the 1D classification, we implemented an early stopping callback with patience of seven epochs to prevent overfitting.

### D. PERFORMANCE EVALUATION

We conducted a 10-fold cross-validation for all experiments, and we present the results in the following format: mean (std). To prevent data leakage, we ensured that exams from the same patient did not appear in different partitions of the cross-validation protocol. In order to evaluate the performance of the employed models, we considered five distinct metrics, including: Sensitivity (Se), Specificity (Spe), F1-score (F1), Area Under Receiver Operating Characteristic curve (AUROC) and Accuracy (Acc). Given the significant class imbalance within the dataset, we defined our best model based on the F1-score.

### E. EXPERIMENTAL SETUP

We conducted experiments to assess the performance and generalizability of our models utilizing distinct approaches: 1D and image-based ECGs. To evaluate the effectiveness of our models, we employed various evaluation setups. Our goal was to assess the overall performance of the models and their ability to generalize to external datasets. The experiments were carried out for both the 1D and image-based ECGs, utilizing the following setups:

1. **Setup 1:**
  a. *Train / Validation / Test (10-fold):* CinC2021 and InCor-DB.
  b. External Validation: None.
2. **Setup 2:**
  a. *Train / Validation / Test (10-fold):* CinC2021.
  b. External Validation: InCor-DB.
3. **Setup 3:**
  a. *Train / Validation / Test (10-fold):* InCor-DB.
  b. *External Validation:* CinC2021.
4. **Setup 4:**
  a. *Train / Validation / Test (10-fold):* InCor-DB.
  b. *External Validation:* CHAPMAN, CPSC, CPSC (extra), GA, NINGBO, PTB-XL.

## III. RESULTS

### A. ONE-DIMENSIONAL-BASED CLASSIFICATION

Table 2 displays performance results for seven proposed architectures, considering AFlut as the positive class. We employed **Setup 1** (aforementioned), utilizing both the CinC2021 and InCor-DB datasets for training, validation, and testing.

**TABLE 2.** Performance results of seven proposed architectures for 1D input data.

| Architectures | Acc | AUROC | F1 | Spe | Se |
| --- | --- | --- | --- | --- | --- |
| *LiteVGG-11* | **77.91** (± 1.73) | **87.17** (± 1.29) | **76.59** (± 1.90) | **71.69** (± 4.73) | **86.53** (± 5.33) |
| *LiteResNet-18* | 76.88 (± 1.61) | 86.64 (± 0.77) | 73.95 (± 2.76) | 75.30 (± 6.66) | 79.06 (± 8.33) |
| *MobileNet* | 77.38 (± 6.67) | 89.52 (± 0.78) | 76.55 (± 4.74) | 70.10 (± 16.97) | 87.53 (± 12.25) |
| *ResNet-50* | 61.51 (± 11.42) | 76.81 (± 9.93) | 64.35 (± 5.78) | 46.73 (± 28.84) | 82.07 (± 17.11) |
| *VGG-16* | 66.49 (± 2.04) | 71.07 (± 2.76) | 59.73 (± 2.77) | 71.48 (± 3.58) | 59.51 (± 4.39) |
| *DenseNet-121* | 75.90 (± 0.84) | 84.46 (± 0.92) | 71.78 (± 1.31) | 77.69 (± 3.64) | 73.39 (± 4.17) |
| *EfficientNet-B2* | 52.49 (± 6.92) | 54.27 (± 2.78) | 28.48 (± 28.55) | 59.28 (± 44.05) | 43.30 (± 45.59) |

In addition to these results, we adopted different strategies: (**Setup 2**) Training/validating/testing exclusively with CinC2021 followed by external validation with InCor-DB (Table 3); and the opposite (**Setup 3**) training with InCor-DB and external validation using CinC2021 (Table 4).

**TABLE 3.** Performance results of seven proposed architectures for one-dimensional input data. Train = CinC2021 dataset. External Validation = InCor-DB dataset.

| Test on CinC2021 dataset |  |  |  |  |  |
| --- | --- | --- | --- | --- | --- |
| Architectures | Acc | AUROC | F1 | Spe | Se |
| *LiteVGG-11* | 72.24 (± 2.13) | 77.43 (± 1.38) | 78.21 (± 2.95) | 57.02 (± 7.39) | 81.73 (± 7.25) |
| *LiteResNet-18* | 70.15 (± 5.05) | 75.85 (± 2.19) | 75.98 (± 8.50) | 53.49 (± 14.11) | 80.64 (± 15.41) |
| *MobileNet* | **71.01** (± 4.54) | **76.49** (± 1.69) | **78.44** (± 3.92) | **45.58** (± 22.44) | **86.90** (± 12.13) |
| *ResNet-50* | 64.63 (± 9.51) | 71.08 (± 3.71) | 67.62 (± 22.32) | 49.25 (± 26.80) | 74.24 (± 29.69) |
| *VGG-16* | 53.45 (± 9.89) | 55.94 (± 4.73) | 48.18 (± 31.63) | 57.28 (± 30.64) | 50.96 (± 34.33) |
| *DenseNet-121* | 67.12 (± 1.38) | 72.52 (± 1.29) | 73.01 (± 2.20) | 58.30 (± 5.75) | 72.63 (± 5.36) |
| *EfficientNet-B2* | 48.34 (± 10.84) | 50.62 (± 2.02) | 34.21 (± 34.65) | 59.46 (± 45.51) | 41.25 (± 45.96) |
| External Validation on InCor-DB dataset |  |  |  |  |  |
| Architectures | Acc | AUROC | F1 | Spe | Se |
| *LiteVGG-11* | 48.93 (± 10.59) | 83.37 (± 3.67) | 32.98 (± 4.20) | 42.56 (± 12.79) | 88.95 (± 4.66) |
| *LiteResNet-18* | 45.25 (± 14.84) | 86.03 (± 3.11) | 32.92 (± 6.19) | 37.80 (± 18.58) | 92.02 (± 9.55) |
| *MobileNet* | 38.04 (± 19.33) | 74.13 (± 4.20) | 29.09 (± 4.39) | 30.16 (± 24.27) | 87.50 (± 12.30) |
| *ResNet-50* | 42.42 (± 25.60) | 66.73 (± 8.37) | 26.20 (± 9.99) | 36.90 (± 33.70) | 77.08 (± 28.61) |
| *VGG-16* | 55.35 (± 22.09) | 54.69 (± 3.11) | 18.07 (± 11.87) | 56.04 (± 31.05) | 51.05 (± 34.61) |
| *DenseNet-121* | 58.30 (± 6.89) | 77.67 (± 2.59) | 35.45 (± 3.41) | 54.52 (± 8.28) | 82.02 (± 3.01) |
| *EfficientNet-B2* | 56.94 (± 33.60) | 50.53 (± 1.29) | 11.95 (± 10.38) | 59.54 (± 46.38) | 40.65 (± 46.62) |

**TABLE 4.** Performance results of seven proposed architectures for one-dimensional input data. Train = InCor-DB dataset. External Validation = CinC2021 dataset.

| Test on InCor-DB dataset |  |  |  |  |  |
| --- | --- | --- | --- | --- | --- |
| Architectures | Acc | AUROC | F1 | Spe | Se |
| *LiteVGG-11* | **95.50** (± 0.99) | **98.33** (± 0.65) | **84.77** (± 3.28) | **96.07** (± 1.08) | **91.70** (± 2.70) |
| *LiteResNet-18* | 95.52 (± 1.10) | 98.28 (± 0.64) | 84.66 (± 4.19) | 96.23 (± 1.58) | 91.26 (± 3.61) |
| *MobileNet* | 87.38 (± 11.42) | 97.62 (± 1.03) | 71.31 (± 14.94) | 86.40 (± 14.02) | 93.53 (± 4.79) |
| *ResNet-50* | 82.93 (± 22.41) | 96.86 (± 2.36) | 69.01 (± 19.15) | 81.84 (± 27.50) | 88.87 (± 13.93) |
| *VGG-16* | 83.88 (± 24.46) | 82.12 (± 20.69) | 56.70 (± 33.21) | 87.72 (± 29.32) | 64.64 (± 33.53) |
| *DenseNet-121* | 93.87 (± 3.76) | 97.07 (± 1.64) | 80.80 (± 7.54) | 94.76 (± 4.54) | 88.29 (± 5.58) |
| *EfficientNet-B2* | 69.69 (± 33.03) | 73.58 (± 23.25) | 48.56 (± 33.66) | 69.47 (± 39.74) | 72.08 (± 34.73) |
| External Validation on CinC2021 dataset |  |  |  |  |  |
| Architectures | Acc | AUROC | F1 | Spe | Se |
| *LiteVGG-11* | 47.56 (± 0.56) | 55.90 (± 1.01) | 31.09 (± 2.12) | 92.78 (± 1.34) | 19.27 (± 1.70) |
| *LiteResNet-18* | 46.89 (± 0.85) | 55.90 (± 0.65) | 28.80 (± 3.44) | 93.78 (± 2.15) | 17.56 (± 2.70) |
| *MobileNet* | 48.66 (± 2.88) | 53.58 (± 1.66) | 38.37 (± 12.07) | 81.07 (± 16.31) | 28.38 (± 14.83) |
| *ResNet-50* | 49.18 (± 4.77) | 55.98 (± 1.18) | 37.95 (± 17.22) | 78.57 (± 27.71) | 30.80 (± 24.83) |
| *VGG-16* | 45.40 (± 6.03) | 54.11 (± 2.79) | 24.21 (± 20.07) | 86.28 (± 28.93) | 19.82 (± 27.44) |
| *DenseNet-121* | 47.68 (± 1.21) | 57.04 (± 0.53) | 31.57 (± 5.40) | 92.07 (± 4.47) | 19.91 (± 4.63) |
| *EfficientNet-B2* | 49.16 (± 7.30) | 52.68 (± 3.04) | 38.59 (± 24.52) | 67.61 (± 39.09) | 37.61 (± 35.87) |

Moreover, Figure 3 presents the accuracy results for the top-performing 1D-based classification model, LiteVGG-11, trained exclusively on InCor-DB and validated externally on individual CinC2021 datasets (**Setup 4**).

**FIGURE 3.**
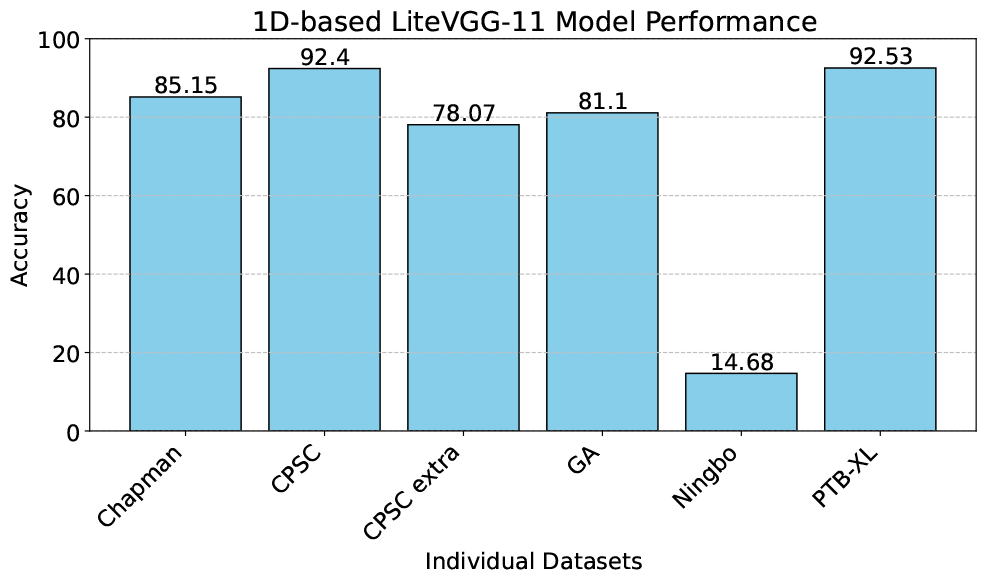
Performance results of LiteVGG-11 one-dimensional-based model trained on InCor-DB dataset and with external validation on each individual CinC2021 dataset.

### B. IMAGE-BASED CLASSIFICATION

Table 5 showcases performance results for five proposed architectures in image-based AFib and AFlut classification. We employed **Setup 1**, utilizing both CinC2021 and InCor-DB for training, validation, and testing.

**TABLE 5.** Performance results of five proposed architectures for image input data.

| Architectures | Acc | AUROC | F1 | Spe | Se |
| --- | --- | --- | --- | --- | --- |
| *MobileNet* | 54.94 (± 13.94) | 76.02 (± 10.99) | 64.16 (± 5.56) | 27.64 (± 31.50) | 92.96 (± 11.62) |
| *ResNet-50* | 49.11 (± 9.51) | 72.35 (± 10.15) | 51.51 (± 19.18) | 26.32 (± 40.88) | 80.11 (± 37.90) |
| *VGG16* | 46.45 (± 7.76) | 50.0 (± 0.0) | 41.16 (± 28.40) | 30.0 (± 48.30) | 70.0 (± 48.30) |
| *DenseNet* | 48.0 (± 9.89) | 64.20 (± 12.53) | 45.65 (± 24.55) | 29.80 (± 47.84) | 72.76 (± 44.76) |
| ***EfficientNet-B2*** | **75.20 (± 3.38)** | **85.50 (± 1.14)** | **71.59 (± 3.66)** | **74.76 (± 13.85)** | **75.74 (± 13.85)** |

Similar to 1D-based classification, we implemented the following strategies: (**Setup 2**) Training/validating/testing exclusively with CinC2021, followed by external validation with InCor-DB (Table 6); and the opposite (**Setup 3**) training with InCor-DB, externally validating using CinC2021 (Table 7).

**TABLE 6.** Performance results of five proposed architectures for image input data. Train = CinC2021 dataset. External Validation = InCor-DB dataset.

| Test on CinC2021 dataset |  |  |  |  |  |
| --- | --- | --- | --- | --- | --- |
| Architectures | Acc | AUROC | F1 | Spe | Se |
| MobileNet | 56.11 (± 11.53) | 67.49 (± 12.28) | 52.94 (± 31.86) | 50.82 (± 45.52) | 59.66 (± 42.85) |
| ResNet-50 | 52.13 (± 11.56) | 68.60 (± 9.97) | 43.22 (± 35.07) | 55.27 (± 48.75) | 50.45 (± 47.51) |
| VGG-16 | 52.25 (± 11.91) | 50.0 (± 0.0) | 45.68 (± 39.31) | 40.0 (± 51.63) | 60.0 (± 51.63) |
| DenseNet-121 | 51.0 (± 13.32) | 65.02 (± 15.25) | 37.50 (± 39.27) | 60.81 (± 46.79) | 44.91 (± 48.58) |
| EfficientNet-B2 | **63.42 (± 3.73)** | **65.08 (± 10.09)** | **74.88 (± 6.0)** | **18.31 (± 29.54)** | **91.85 (± 18.04)** |
| External Validation on InCor-DB dataset |  |  |  |  |  |
| Architectures | Acc | AUROC | F1 | Spe | Se |
| MobileNet | 45.53 (± 32.33) | 65.98 (± 20.96) | 19.31 (± 13.19) | 42.65 (± 43.18) | 63.64 (± 46.31) |
| ResNet-50 | 52.15 (± 36.67) | 65.88 (± 14.41) | 21.29 (± 13.43) | 51.72 (± 49.46) | 54.90 (± 44.87) |
| VGG-16 | 42.74 (± 37.45) | 50.0 (± 0.0) | 14.49 (± 12.47) | 40.0 (± 51.63) | 60.0 (± 51.63) |
| DenseNet-121 | 52.58 (± 36.16) | 61.18 (± 16.95) | 11.99 (± 12.43) | 53.61 (± 49.75) | 46.12 (± 49.20) |
| EfficientNet-B2 | 20.84 (± 14.84) | 68.79 (± 14.27) | 25.53 (± 2.91) | 8.77 (± 18.41) | 96.63 (± 7.70) |

**TABLE 7.** Performance results of five proposed architectures for image input data. Train = InCor-DB dataset. External Validation = CinC2021 dataset.

| Test on InCor-DB dataset |  |  |  |  |  |
| --- | --- | --- | --- | --- | --- |
| Architectures | Acc | AUROC | F1 | Spe | Se |
| MobileNet | 91.19 (± 6.74) | 97.71 (± 1.84) | 74.92 (± 11.39) | 91.57 (± 9.11) | 87.54 (± 17.27) |
| ResNet-50 | 72.37 (± 32.93) | 92.51 (± 15.03) | 60.81 (± 24.52) | 69.39 (± 39.36) | 91.99 (± 14.96) |
| VGG-16 | 71.19 (± 31.06) | 50.0 (± 0.0) | 4.39 (± 9.27) | 80.0 (± 42.16) | 20.0 (± 42.16) |
| DenseNet-121 | 40.81 (± 35.60) | 85.79 (± 19.38) | 34.17 (± 25.11) | 33.42 (± 87.86) | 87.86 (± 30.17) |
| EfficientNet-B2 | **94.08 (± 2.59)** | **97.03 (± 2.41)** | **80.11 (± 8.57)** | **95.06 (± 3.46)** | **87.76 (± 11.09)** |
| External Validation on CinC2021 dataset |  |  |  |  |  |
| Architectures | Acc | AUROC | F1 | Spe | Se |
| MobileNet | 82.66 (± 8.96) | 84.96 (± 3.19) | 46.97 (± 12.03) | 85.71 (± 13.11) | 61.58 (± 25.22) |
| ResNet-50 | 51.71 (± 5.52) | 52.51 (± 5.87) | 48.30 (± 19.72) | 60.56 (± 39.63) | 46.18 (± 33.05) |
| VGG-16 | 43.09 (± 9.71) | 50.0 (± 0.0) | 15.23 (± 32.11) | 80.0 (± 42.16) | 20.0 (± 42.16) |
| DenseNet-121 | 55.70 (± 8.45) | 53.58 (± 13.65) | 59.41 (± 27.17) | 29.04 (± 42.42) | 72.37 (± 40.12) |
| EfficientNet-B2 | 47.26 (± 2.06) | 54.64 (± 2.94) | 32.28 (± 8.62) | 89.13 (± 7.40) | 21.06 (± 7.10) |

Figure 4 depicts accuracy results for our top-performing image-based classification model, EfficientNet-B2. This model was solely trained on InCor-DB and externally validated on individual CinC2021 datasets (**Setup 4**).

**FIGURE 4.**
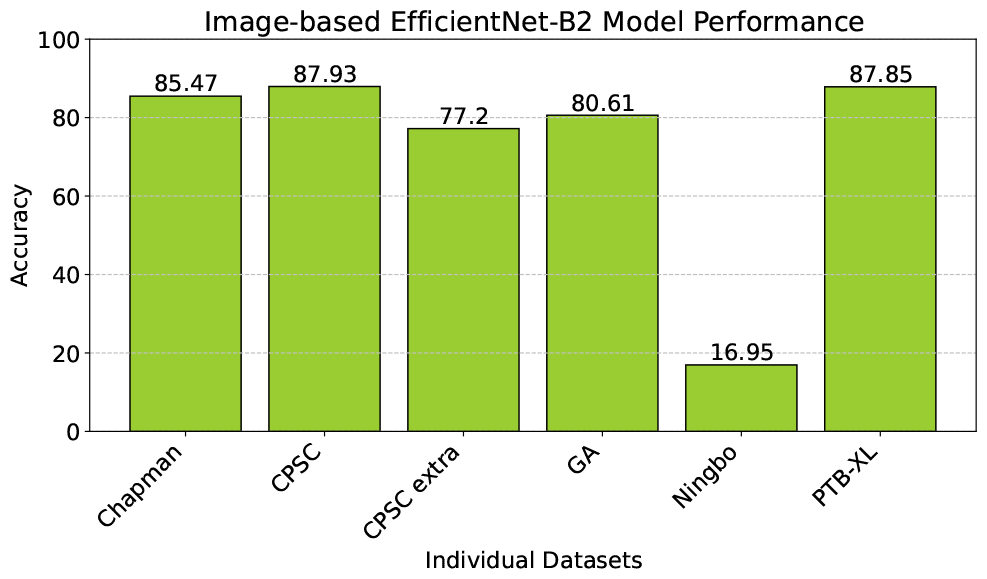
Performance results of EfficientNet-B2 image-based model trained on InCor-DB dataset and with external validation on each individual CinC2021 dataset.

## IV. DISCUSSION

In our current study, we employed different approaches to evaluate the potential of CNN models in distinguishing between AFib and AFlut. As far as we know, we are the first to present an assessment of AFib and AFlut specific discrimination using end-to-end CNNs, demonstrating the feasibility of reasonably distinguishing these two diagnoses.

Our primary findings can be summarized as follows and will be further discussed bellow: (1) When utilizing all available databases (CinC2021 and InCor-DB), only models based on 1D data achieved the capacity to discriminate between AFib and AFlut, exhibiting reasonable performance; (2) Models based on 2D data demonstrated poor performance, with the exception of the EfficientNet-B2 model; (3) Concerning the available datasets, models trained solely on the CinC2021 databases struggle to differentiate the study classes, resulting in metrics that closely resemble chance levels. Conversely, models exclusively based on the InCor-DB private dataset successfully separated the classes; (4) We emphasize the significance of evaluating the separability of classes within the study dataset before contemplating the combination of AFib and AFlut exams into a single class for further analysis. In our research, we observed a clear differentiation between these two classes in the InCor-DB dataset, but this was not evident in the case of the CinC2021 databases; Additionally, (5) Concerning the CinC2021 datasets, we advise exercising caution when using the Ningbo dataset. Our results indicate that a majority of the exams labeled as AFlut are predicted as AFib by our models.

### A. MODELS BASED ON BOTH CINC2021 AND INCOR-DB DATASET

In 1D models (Table 2), EfficientNet-B2 struggled to address the problem, performing close to chance level, while LiteVGG-11 had the best performance. Moreover, among image-based models (Table 5), performance was generally poor, except for EfficientNet-B2, which exhibited results similar to 1D models.

Previous research aimed at distinguishing AFib and AFlut using MIT-BIH datasets [28] that has limitations due to a limited number of subjects in long-term Holter recordings [12]. These recordings failed in representing arrhythmia diversity compared to CinC2021 databases and the InCor-DB dataset, which feature more subjects and exams.

### B. MODELS BASED ON CINC2021 DATASET, WITH EXTERNAL VALIDATION ON INCOR-DB DATASET

The results from the CinC2021 databases indicated that our proposed image-based networks struggled to differentiate AFib from AFlut, with most evaluation metrics hovering near chance level. Table 6 supports this assessment, particularly for the CinC2021 test set. Even the EfficientNet-B2 architecture, while showing better performance compared to others, seemed to assign exams predominantly to one class, as indicated by specificity and sensitivity metrics. In contrast, our 1D-based models (Table 3) performed reasonably well, particularly LiteVGG-11, LiteResNet-18, and MobileNet architectures.

Upon analyzing data distribution across individual datasets within the CinC2021 database, we observed that the imbalanced data can potentially be leading our models to learn to differentiate datasets rather than addressing the primary task of discriminating AFib and AFlut. As shown in Table 1, the CinC2021 database comprises 5,238 AFib exams and 8,373 AFlut exams. Given that a significant portion of AFlut samples originate from the Ningbo First Hospital 12-lead ECG dataset, it’s reasonable that our models are distinguishing exams based on their origin rather than their underlying arrhythmia type.

### C. MODELS BASED ON INCOR-DB DATASET, WITH EXTERNAL VALIDATION ON CINC2021 DATASET

When utilizing the InCor-DB private dataset (Tables 4 and 7), despite class imbalance, our models achieved outstanding performance, with Acc and AUROC scores exceeding 90%. However, during external validation on the CinC2021 dataset, our models struggled to generalize. Given these findings, it appears appropriate to merge AFib and AFlut labeled exams in future ECG classification experiments using the CinC2021 databases, as our models’ performance approaches chance level, contrasting with the clear discriminability of the two classes in the InCor-DB dataset.

Considering the significant class imbalance, we evaluated model performance based on F1-score, selecting MobileNet for 1D models and EfficientNet-B2 for image-based models as our top-performing models. These models were exclusively trained on the InCor-DB dataset and validated externally with each dataset from the CinC2021 databases. In most cases, our trained models performed well, except for the CPSC and Ningbo datasets (Table 4).

For the CPSC dataset, most exams were predicted as AFib, which aligns with all exams being labeled as such. However, regarding the Ningbo dataset, where AFlut is the designated label for all exams, our model predominantly predicted AFib for exams labeled as AFlut. This discrepancy may be attributed to potential variations in diagnostic criteria among cardiologists from different nations when distinguishing AFib and AFlut. Conversely, diagnoses in the InCor-DB dataset originated from cardiologists within the same hospital, likely following consistent diagnostic criteria. In the worst-case scenario, one might argue that exams in the Ningbo dataset could be mislabeled or, more concerning, misdiagnosed.

## V. CONCLUSION

In this work we explored the potential of CNN models to distinguish between AFib and AFlut based on ECG data. Contrary to our initial expectations, our findings suggest that one-dimensional models generally outperformed image-based models in this discrimination task. This discrepancy underscores the complexity of translating clinical intuition into computational models and highlights the importance of empirical validation in machine learning research. Specifically, our analysis demonstrated that 1D models exhibited superior performance, particularly when trained on the InCor-DB dataset. However, model performance decreased when validated on CinC2021 dataset. Additionally, we emphasized the importance of careful dataset selection and evaluation, as well as consistency in exam labeling. While models trained on InCor-DB achieved high accuracy, there were discrepancies in model predictions for the Ningbo dataset, highlighting the need for standardized diagnostic criteria. Our research suggests that by overcoming these challenges, CNN models can enhance arrhythmia diagnosis, benefiting healthcare and clinical outcomes.

## Data Availability

Publicly available datasets were analyzed in this study. This data can be found in the PhysioNet databank: https://physionet.org/content/challenge-2021/1.0.2/. Private InCor-DB dataset will be made available on request.

## ETHICS STATEMENT

This study was approved by the Institutional Review Board (IRB), with registration CAAE 45070821.3.0000.0068, as part of the Machine Learning in Cardiovascular Medicine Project. It is worth noting that this research constitutes a secondary analysis of fully anonymized data stored in the InCor-DB.

## AUTHORS CONTRIBUTIONS STATEMENT

E.R. Conceptualization, Methodology, Implementation and Writting. Q.B.S Conceptualization, Methodology, Implementation and Writting. F.M.D. Conceptualization, Methodology, Implementation and Writting. J.E.K. and M.A.G. Supervision and Review. All authors analyzed the results and revised critically the manuscript. All authors read and approved the submitted manuscript.

## ACKNOWLEDGEMENTS

This study was financially supported in part by São Paulo Research Foundation (FAPESP) – grant n^*o*^ 2021/12935-0, the Foxconn Brazil, and the Zerbini Foundation as part of the research project “Machine Learning in Cardiovascular Medicine”.

---

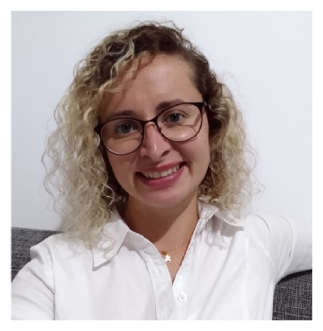

ESTELA RIBEIRO received in 2015 the B. Sc. degree in mechanical engineering from FSA University Center, São Paulo, Brazil. Obtained the M.Sc. and the ph.D. degrees in electrical engineering from FEI University Center, São Paulo, Brazil, in 2017 and 2020, respectively. Her research interests include pattern recognition, cognitive perception, biomedical signal processing and machine learning. She is currently a Researcher at the Laboratory of Biomedical Informatics of the Heart Institute, Clinics Hospital, University of São Paulo Medical School.

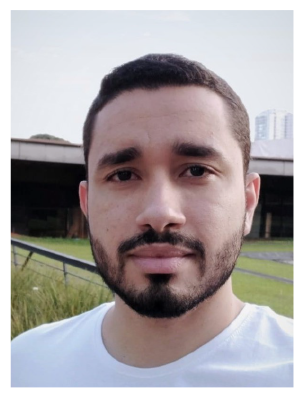

QUENAZ BEZERRA SOARES received the B.Sc. degree in electrical engineering from the Federal University of Viçosa (UFV) in 2021. He is currently pursuing a M.Sc. in biomedical engineering at the University of São Paulo (USP), working with lightweight deep learning applications in wearable electrocardiogram signals. Furthermore, he is currently a Researcher at the Laboratory of Biomedical Informatics of the Heart Institute (InCor), Clinics Hospital, University of São Paulo Medical School. His research interests include biomedical signal processing, machine learning and evolutionary algorithms.

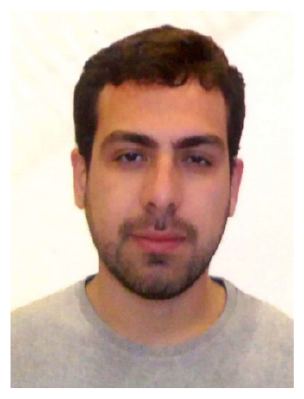

FELIPE MENEGUITTI DIAS received the B.Sc. and the M.Sc. degree from the Federal University of Juiz de Fora (UFJF) in 2017 and 2020, respectively. He is currently pursuing a Ph.D. in biomedical engineering at the University of Sao Paulo (USP), working with machine learning applications in electrocardiogram and photoplethysmogram biomedical signals. Furthermore, he is a researcher at the Heart Institute (Incor-HCFMUSP). His research interests include biomedical signal processing, machine learning, and compressive sensing.

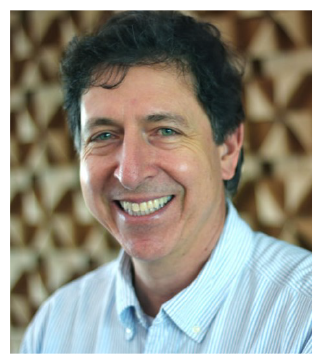

JOSE EDUARDO KRIEGER, MD, PhD, is Professor of Genetics and Molecular Medicine at the University of Sao Paulo Medical School and Director of the Laboratory of Genetics & Molecular Cardiology at the Heart Institute (InCor). His research interests are focused on the genetic determinants of cardiovascular diseases to improve health management algorithms and to the development of novel therapeutics.

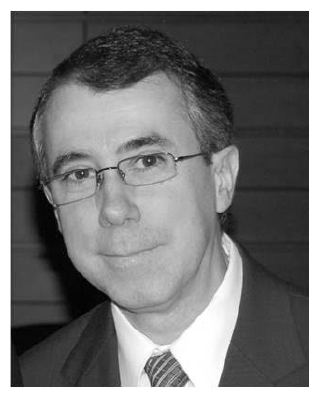

MARCO ANTONIO GUTIERREZ received the B.Eng. and D.Sc. degrees in electrical engineering from the University of São Paulo, Brazil, in 1985 and 1996, respectively. He has been with the Heart Institute, University of São Paulo, since 1986, where he is currently the Head of the Biomedical Informatics Laboratory and the Informatics Division. He is also an Assistant Professor at the Polytechnic School, since 1997, and School of Medicine, since 2004, University of São Paulo, Brazil. His research interests include biomedical image and signal processing and health information systems.

